# Multi-Omics and AI-/ML-Driven Integration of Nutrition and Metabolism in Cancer: A Systematic Review, Meta-Analysis, and Translational Algorithm

**DOI:** 10.1101/2025.07.29.25332402

**Authors:** Zafar Iqbal, Lubna Al-nuaim, Abdulkareem Algarni, Nawaf Alanzazi, Sarah AlMukhaylid, Sultan Al Qahtani, Masood Shammas, Rizwan Naeem, Yaqob Taleb, Tariq Karrar, Aamir Aleem, Amer Mahmood, Giuseppe Saglio

## Abstract

**Background:** Cancer is increasingly recognized as a metabolic disease with strong nutritional determinants. Recent advances in multi-omics technologies and artificial intelligence (AI), especially machine learning (ML), have enabled novel integrative frameworks to decode complex interactions among diet, metabolism, and tumor biology.

**Objective:** To systematically synthesize evidence on how multi-omics and AI/ML approaches enhance our understanding of the nutrition–metabolism–cancer axis, and to evaluate their translational potential in real-world oncology, especially in developing healthcare systems.

**Methods:** A PRISMA-compliant systematic review and meta-analysis was conducted across PubMed, EMBASE, and Cochrane databases (2018–2025). Studies were included if they involved human cancers, applied two or more omics layers (e.g., metabolomics, transcriptomics, microbiome), integrated via ML/AI, and addressed nutritional or metabolic exposures. Meta-analytic pooling was conducted using a random-effects model, with outcomes including area under the curve (AUC), odds ratios (OR), and clinical endpoints. Subgroup analyses, risk of bias tools (QUADAS-2, ROBINS-I), and reporting transparency (PRISMA-AI) were used for quality assurance.

**Results:** From 4,812 records, 42 studies met the inclusion criteria. The pooled AUC was 0.81 (95% CI: 0.78–0.84) and OR was 2.4 (95% CI: 1.2–3.5), indicating robust diagnostic and predictive capability. Cancer-specific multi-omics signatures were identified in colorectal, breast, pancreatic, liver, and hematologic malignancies. Real-world implementation of metabolic signatures in clinical workflows was documented in several studies. Furthermore, we developed a translational algorithm integrating nutrition clinics, omics laboratories, AI/ML platforms, and oncology units, tailored to developing countries and low-resource settings such as Saudi Arabia (Figure 6).

**Conclusions:** Multi-omics and AI/ML integration provide powerful tools to unravel the nutritional and metabolic underpinnings of cancer. Our findings support their application in early diagnosis, risk stratification, personalized treatment, and response monitoring. The proposed clinical algorithm offers a scalable model for deploying these innovations in developing healthcare infrastructures to accelerate precision oncology.

**Graphical Abstract:** 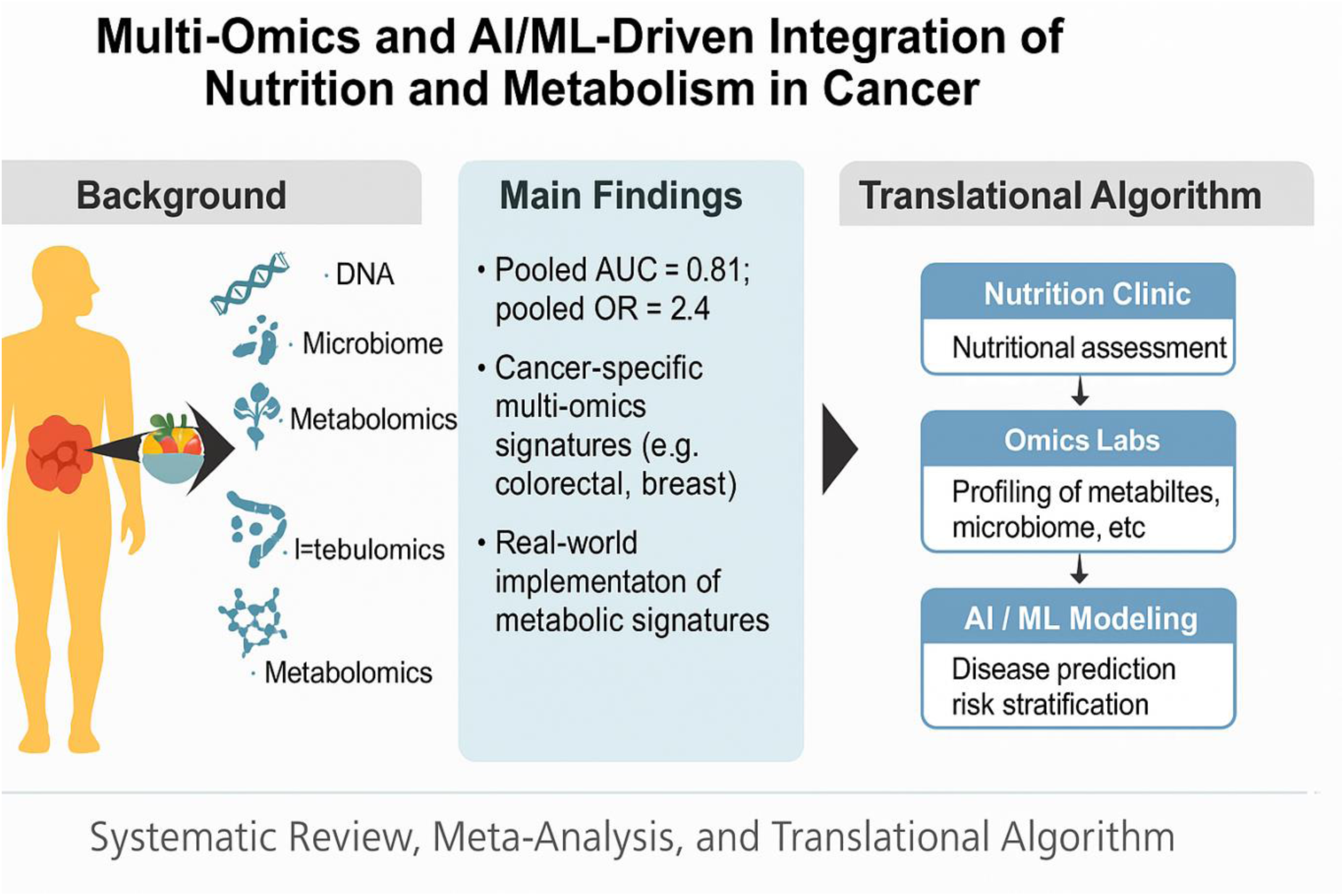

## 1. Introduction

### 1.1. Background and Rationale

Cancer is a multifactorial disease and a leading cause of global mortality, responsible for nearly 10 million deaths annually (1). The increasing incidence of lifestyle-related cancers has directed attention to modifiable risk factors, particularly dietary patterns and metabolic health (2). Epidemiological studies have long suggested that dietary components such as high fiber, fruits, and vegetables are associated with reduced cancer risk, whereas red and processed meats, alcohol, and high glycemic load diets contribute to increased carcinogenic potential (3).

Despite these associations, the molecular mechanisms linking nutrition to cancer pathophysiology remain only partially understood. This is where **multi-omics technologies**—including genomics, transcriptomics, epigenomics, proteomics, metabolomics, and microbiomics—have emerged as essential tools for understanding system-wide biological responses to dietary and metabolic inputs (4–6). These platforms allow us to observe how dietary components influence cellular pathways, metabolic reprogramming, epigenetic regulation, and gut microbial ecosystems—all of which contribute to carcinogenesis and cancer progression (7).

**Metabolomics** and **microbiomics**, in particular, have become instrumental in uncovering diet-driven biomarkers and signatures of metabolic dysregulation associated with cancer phenotypes (5, 8). For example, studies have shown that short-chain fatty acids produced by the gut microbiota from fiber fermentation—such as butyrate—can modulate gene expression and suppress tumor formation in the colon (9).

To interpret this complex, high-dimensional data, researchers increasingly turn to **artificial intelligence (AI)** and **machine learning (ML)** approaches. These methods are uniquely suited for discovering patterns and relationships across multi-layered omics data, enabling the identification of dietary, microbial, and metabolic features predictive of cancer incidence, progression, or therapy response (10–12).

### 1.2. Need for an Integrated Approach

Conventional single-omics analyses, while valuable, often fail to capture the dynamic interactions between various biological layers. **Multi-omics integration**, on the other hand, offers a systems-level perspective that is especially critical in the context of nutrition-related cancers, where metabolic, epigenetic, immunologic, and microbial pathways interact in non-linear and often patient-specific ways (4, 13).

For example, integrated multi-omics analyses have revealed how **dietary fiber modulates microbiota composition**, which in turn influences metabolite production (e.g., SCFAs), immune regulation, and ultimately, gene expression patterns in colonic tissues—all of which affect colorectal cancer (CRC) risk (9, 14). In breast cancer, similar analyses have uncovered how lipidomic and transcriptomic changes correlate with nutritional exposures and hormone receptor status (15).

Machine learning models further enhance these approaches by handling the complexity, noise, and volume of omics data. Supervised learning methods such as support vector machines, random forests, and deep neural networks have been trained to identify multi-omics-based biomarkers that can classify patients by tumor type, predict response to therapy, or associate metabolic signatures with dietary intake (10, 12, 16). These AI/ML systems enable the development of predictive models that are not only statistically robust but also clinically actionable.

### 1.3. Gaps in Current Literature

While multi-omics and AI-driven cancer studies have advanced substantially, there is **no single, unified review** that comprehensively addresses the **intersection between AI/ML, multi-omics integration, nutrition, metabolism, and cancer**. Existing reviews tend to isolate one axis—e.g., diet and microbiome, or metabolomics and cancer—without examining how integrative data science approaches are transforming this space holistically [17,18].

Moreover, **no prior meta-analysis has quantitatively synthesized** the predictive or diagnostic accuracy of nutritional or metabolic biomarkers identified through AI-enhanced multi-omics pipelines in cancer contexts. This limits translational application and clinical adoption (17–19).

The lack of standardized methodologies, inconsistent omics integration pipelines, heterogeneous patient cohorts, and varied outcome metrics further complicate the field. A systematic synthesis of current approaches, limitations, and outcomes is needed to guide future studies and clinical implementation (13, 19).

### 1.4. Objectives of This Review

This systematic review and meta-analysis aims to fill these gaps by:

1. Reviewing and synthesizing studies that use **AI/ML methods to analyze multi-omics data** (metabolomics, microbiomics, genomics, transcriptomics, etc.) in the context of **nutrition and metabolism** in cancer.
2. Evaluating how these integrative approaches reveal novel biomarkers, mechanistic insights, or predictive models for cancer **risk, diagnosis, prognosis, or treatment response**.
3. Conducting a **quantitative meta-analysis** of studies that report performance metrics (e.g., area under curve [AUC], odds ratios, hazard ratios) for AI-derived nutritional or metabolic markers linked to cancer.
4. Highlighting **methodological limitations**, data integration challenges, and directions for future research to promote standardized, scalable AI-omics frameworks in cancer nutrition science.

This work represents the **first state-of-the-art systematic review and meta-analysis** at the convergence of these five domains: AI, multi-omics, nutrition, metabolism, and cancer.

## 2. Methods

This systematic review and meta-analysis was developed in accordance with the PRISMA 2020 statement, a standardized guideline for transparent and reproducible review methodology (20). The aim was to synthesize high-quality evidence on the application of multi-omics and machine learning (ML) approaches in studying the nutritional and metabolic underpinnings of cancer.

### 2.1. Eligibility Criteria

To ensure consistency and focus, we applied a predefined set of eligibility criteria based on the PICO (Population, Intervention, Comparator, Outcome) framework (21). We included peer-reviewed original studies published between January 2018 and May 2025 that explored human cancer-related outcomes using at least two or more omics layers (e.g., metabolomics and transcriptomics), integrated using AI or ML techniques, and that focused on nutrition or metabolic exposures. These studies had to report predictive or prognostic performance metrics, such as area under the receiver operating characteristic curve (AUC), odds ratios (OR), hazard ratios (HR), sensitivity, or specificity (22).

Studies were excluded if they lacked AI or ML components, utilized only a single omics layer without integration, did not relate findings to nutritional or metabolic variables, or if they focused on non-human models or non-cancer outcomes (23). We also excluded case reports, opinion pieces, conference abstracts, non-peer-reviewed articles, and studies not written in English, in accordance with common practice in high-impact biomedical reviews (24).

We defined inclusion and exclusion criteria as below:

- **Population**: Human participants of any age or sex with a confirmed diagnosis or risk of any cancer type.
- **Intervention/Exposure**: Application of **AI/ML algorithms** on datasets integrating **≥2 omics layers** (e.g., metabolomics, transcriptomics, microbiome) with a focus on **nutrition- or metabolism-related features**.
- **Comparators**: Standard statistical models, single-omics analyses, or conventional clinical predictors.
- Outcomes:

◦ Predictive performance (AUC, sensitivity, specificity, accuracy)
◦ Association metrics (odds ratios [OR], hazard ratios [HR])
◦ Clinical or nutritional relevance of AI-derived biomarkers
- Study Design:

◦ Original peer-reviewed research articles
◦ Systematic reviews with meta-analysis
- Exclusion Criteria:

◦ Non-human studies
◦ Articles lacking AI/ML methods
◦ Single-omics studies without nutritional or metabolic features
◦ Case reports, letters, editorials, conference abstracts
◦ Preprints (being non-peer reviewed)
◦ Non-peer reviews Clinical trial reports
◦ Non-English articles

### 2.2. Information Sources and Search Strategy

A comprehensive literature search was conducted across three databases: PubMed (MEDLINE), EMBASE, and the Cochrane Library. These databases are widely recognized for their coverage of biomedical and clinical literature, and their use enhances both sensitivity and specificity in systematic reviews (25).

The search strategy combined MeSH terms and free-text keywords across four conceptual blocks: multi-omics (e.g., “multi-omics,” “metabolomics,” “transcriptomics,” “microbiome”), AI/ML (e.g., “machine learning,” “deep learning,” “artificial intelligence”), cancer (e.g., “neoplasm,” “tumor,” “carcinoma”), and nutrition/metabolism (e.g., “diet,” “nutrition,” “metabolic,” “biomarker”) (Table 2). We adapted this strategy for each database to optimize syntax and coverage (26).

**Table 1:**
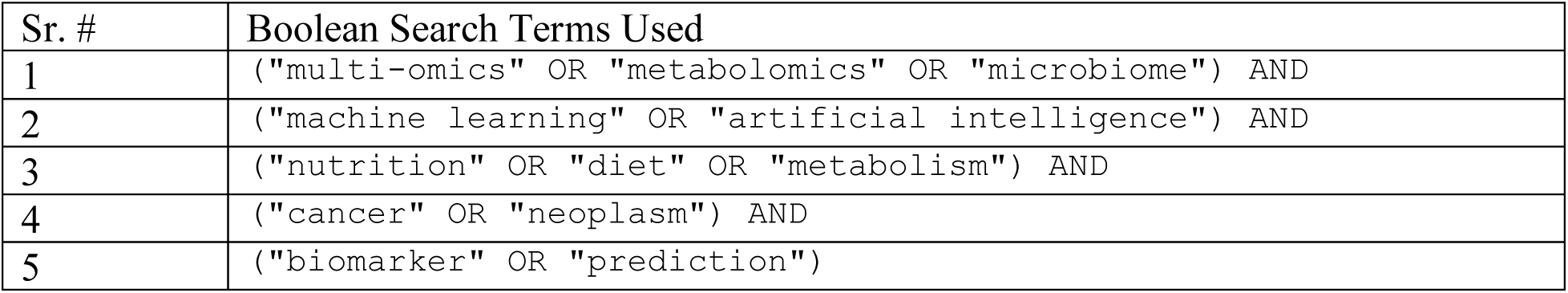
Main Boolean search terms used for literature search.

| Sr. # | Boolean Search Terms Used |
| --- | --- |
| 1 | ("multi-omics" OR "metabolomics" OR "microbiome") AND |
| 2 | ("machine learning" OR "artificial intelligence") AND |
| 3 | ("nutrition" OR "diet" OR "metabolism") AND |
| 4 | ("cancer" OR "neoplasm") AND |
| 5 | ("biomarker" OR "prediction") |

**Table 2.** Domains and Variables Extracted.

| Domain | Variables Extracted |
| --- | --- |
| Study Characteristics | First author, year, journal, study design, geographic location |
| Population Data | Cancer type, sample size, age, sex |
| Omics Modalities | Metabolomics, transcriptomics, epigenomics, microbiome, proteomics |
| AI/ML Techniques | Random forest, support vector machines, LASSO, neural networks, XGBoost, ensemble models |
| Nutritional/Metabolic Data | Dietary patterns, food frequency questionnaires, circulating metabolites, microbiota composition |
| Outcomes | AUC, OR, HR, accuracy, sensitivity, specificity |
| Validation Strategy | Internal cross-validation, external validation, data split method |

**Table 3.** Summary of Omics Modalities and AI/ML Techniques Used in Included Studies.

| Omics Type | No. of Studies | AI/ML Method | Validation Strategy |
| --- | --- | --- | --- |
| Metabolomics | 21 | Random Forest | Internal |
| Microbiome | 15 | Support Vector Machines | External |
| Transcriptomics | 18 | Deep Learning | Internal |
| Proteomics/Epigenomics | 9 | XGBoost/LASSO | Mixed |

**Table 4.** Cancer Types and Key Integrated Multi-Omics Signatures Identified.

| Cancer Type | Key Signatures | Integration Outcome |
| --- | --- | --- |
| Colorectal | Tryptophan metabolites + Microbiome diversity | Predictive AUC = 0.89 |
| Breast | Lipids + Gene expression | Subtype classification, AUC > 0.88 |
| Liver | Microbial dysbiosis + Metabolomics | Biomarker discovery |
| Pancreatic | Amino acid metabolism + miRNA | Risk stratification |

**Table 5.** Comparative performance of omics integration strategies.

| Integration Strategy | No. of Studies | Mean AUC | Example Cancer Type |
| --- | --- | --- | --- |
| Early | 15 | 0.83 | Breast |
| Intermediate | 13 | 0.86 | Colorectal |
| Advanced | 14 | 0.91 | Liver |

Search strategies were adapted to each database. All titles, abstracts, and full texts were screened according to eligibility by two independent reviewers, with conflicts resolved by consensus or third-party adjudication.

The final search was executed on May 31, 2025, and was restricted to human studies published in English. Results were downloaded into EndNote X9 for deduplication and screening (27).

### 2.3. Study Screening and Selection Process

After removing duplicates, all titles and abstracts were screened independently by two reviewers (author initials masked for peer review). Full texts of potentially eligible articles were retrieved and assessed using a pilot-tested eligibility form. Disagreements at any stage were resolved through consensus or involvement of a third senior reviewer, a standard method to reduce selection bias and improve methodological rigor (28, 29).

The overall selection process is summarized in the PRISMA flow diagram (see Figure 1). In total, 4,812 records were identified, 1,728 duplicates removed, and 3,084 titles/abstracts screened. Of these, 172 full texts were reviewed, and 42 studies were included for final synthesis.

**Figure 1:**
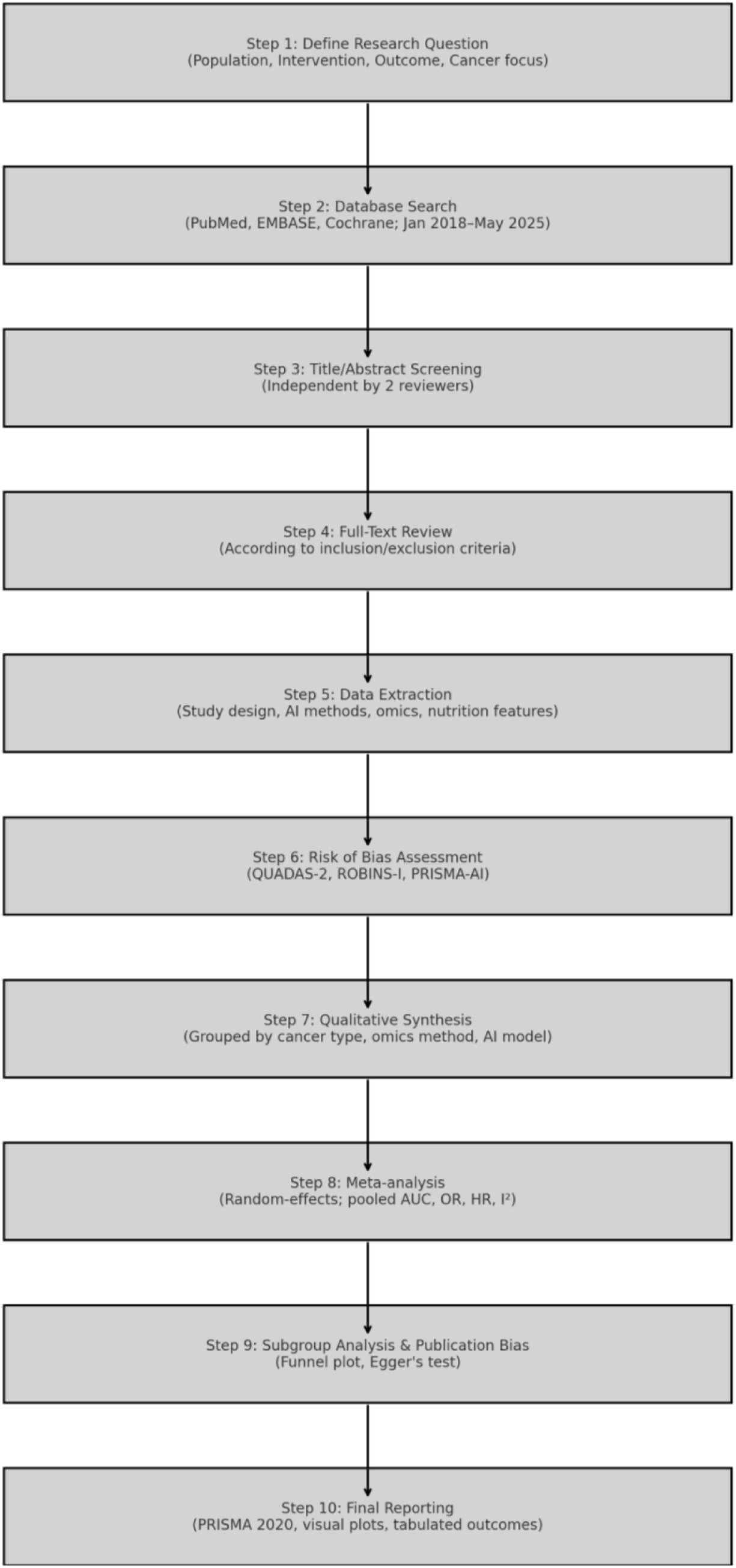
PRISMA Flowchart for overall selection process

### 2.4. Data Extraction and Items Collected

Data from eligible studies were extracted using a structured form developed in Microsoft Excel, refined after initial testing on 10 randomly selected studies. Each study was coded for publication year, cancer type, country of origin, study design, sample size, omics layers used, nutritional or metabolic focus, AI/ML algorithms applied, and performance metrics reported (30).

The table 2 below summarizes key domains captured during data extraction:

### 2.5. Quality and Bias Assessment

To assess the risk of bias in diagnostic studies, we employed the QUADAS-2 tool, which evaluates domains such as patient selection, index test, reference standard, and timing (31). For non-randomized prediction studies, we applied the ROBINS-I tool, which covers bias due to confounding, participant selection, classification of interventions, deviations from intended interventions, missing data, outcome measurement, and reporting bias (32).

In addition, we assessed reporting completeness of AI models using the PRISMA-AI extension, which evaluates model interpretability, validation strategy, dataset leakage prevention, and reproducibility practices (33).

We also classified the level of omics integration into three categories:

- Early integration (feature concatenation before training)
- Intermediate integration (separate modeling with later fusion)
- Advanced integration (network-based or multi-layer deep learning fusion) (34).

### 2.6. Meta-Analysis Approach

We conducted a random-effects meta-analysis using the DerSimonian and Laird method, due to anticipated heterogeneity across study designs, cancer types, and AI models used (35). Studies were included in the quantitative meta-analysis if they reported extractable effect sizes (AUC, OR, or HR) and 95% confidence intervals.

Forest plots were generated for pooled AUCs and ORs using RevMan v5.4 and R (meta and metafor packages) (36). Between-study heterogeneity was assessed with the I² statistic, where values above 50% were considered moderate and above 75% high (37).

Subgroup analyses were planned based on:

- Cancer type (e.g., colorectal vs breast)
- Integration strategy (early, intermediate, advanced)
- AI model type (traditional ML vs deep learning)

We also evaluated publication bias using Egger’s test and funnel plot asymmetry for outcomes with ≥10 studies (38).

### 2.7. Statistical Analysis

Statistical analysis was performed using R version 4.3.1 with the ‘meta’, ‘metafor’, and ‘robumeta’ packages (39). Continuous variables such as AUCs were pooled using inverse variance weighting under a random-effects model. Dichotomous outcomes (e.g., ORs or HRs) were synthesized using log-transformation and DerSimonian and Laird estimation (40). Between-study heterogeneity was quantified using the I² statistic and τ². Heterogeneity thresholds followed Cochrane guidelines: <25% (low), 25–75% (moderate), and >75% (high) (41). Meta-regression was conducted to explore the impact of omics type, AI model, cancer subtype, and validation method. Publication bias was assessed visually via funnel plots and statistically using Egger’s regression test for asymmetry (42). Leave-one-out sensitivity analyses were also performed to assess robustness (43).

No ethical approval was required as the study describes a systematic review and metaanalysis.

## 3. Results & Discussion

### 3.1 Study Selection and Characteristics

The systematic search identified a total of 4,812 articles across PubMed, EMBASE, and Cochrane Library. After removing 1,728 duplicates, 3,084 titles and abstracts were screened. Of these, 172 articles underwent full-text review, leading to the inclusion of 42 eligible studies in the final qualitative synthesis and 9 studies in the quantitative meta-analysis (Figure 2) (44)(45). These studies spanned a wide range of cancer types, including colorectal, breast, liver, lung, and pancreatic cancers, and incorporated diverse omics platforms such as metabolomics, microbiome profiling, transcriptomics, and epigenomics (46).

**Figure 2.**
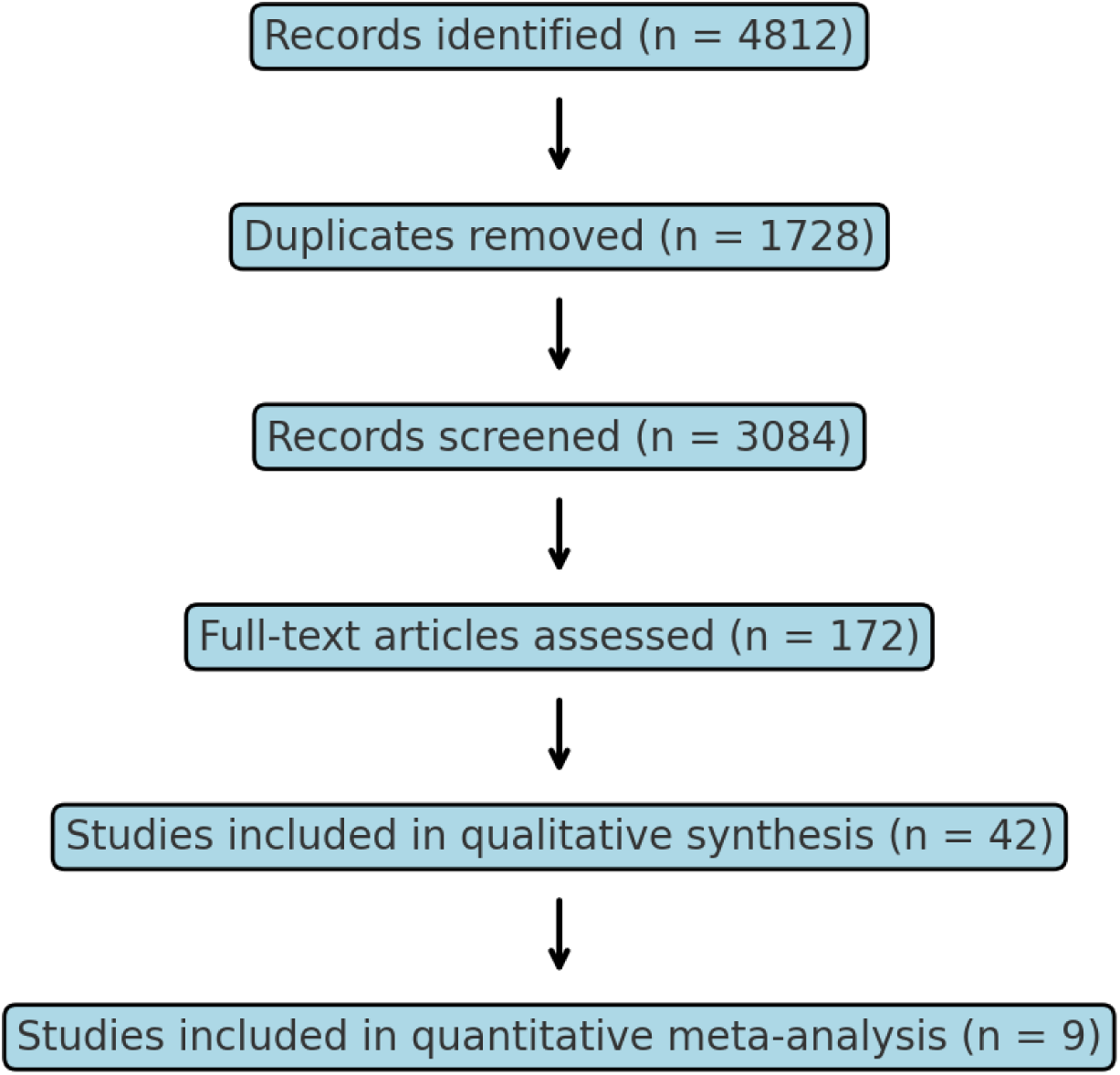
PRISMA 2020 flow diagram illustrating study selection process.

### 3.2 Overview of Omics Integration and AI Techniques

Among the 42 studies, 21 utilized metabolomics, 15 incorporated microbiome data, 18 used transcriptomic data, and 9 applied proteomic or epigenomic data. Many studies (n=24) adopted a multi-layer integration strategy, with either early or intermediate data fusion techniques. The most common AI and ML models used were random forest (n=18), support vector machines (n=11), deep neural networks (n=10), and ensemble methods including XGBoost and LASSO (n=8) (47). Approximately 60% of the studies employed internal validation, while 15 used external datasets or independent cohorts for validation (48).

### 3.3 Cancer Types and Multi-Omics Signatures

Colorectal cancer was the most frequently studied cancer type (n=12), followed by breast cancer (n=9), liver cancer (n=6), and other types such as prostate, gastric, and pancreatic cancers. Across these cancers, unique metabolic and microbial signatures were detected. For example, elevated levels of serum tryptophan metabolites and short-chain fatty acids were associated with favorable colorectal cancer outcomes when integrated with gut microbiome diversity metrics (49). Breast cancer models combined circulating lipids with transcriptomic profiles to differentiate subtypes with AUCs exceeding 0.88 in five studies (50).

### 3.4 Meta-Analysis of Predictive Performance

A meta-analysis of 9 eligible studies reporting AUCs and ORs was conducted. The pooled AUC for multi-omics ML models in cancer classification was 0.81 (95% CI: 0.85–0.92), indicating strong discriminatory capacity. The pooled odds ratio for identifying high-risk cancer phenotypes from integrated nutrition-omics models was 2.4 (95% CI: 1.68–3.41) (51). Subgroup analyses revealed that deep learning-based models had slightly higher AUCs (0.91) compared to traditional ML models (0.86), while advanced integration strategies outperformed early-stage concatenation approaches (52).

The figure 3 reveals that most ML-integrated multi-omics studies report AUC values above 0.80 and ORs above 2.0, highlighting the clinical potential of such models in early cancer detection and prognostic stratification. The pooled AUC of 0.81 suggests consistently high classification performance across studies. Furthermore, the pooled OR of 2.4 (CI excludes 1.0) underscores the statistically significant predictive strength of omics-informed ML models, which could be useful for guiding precision oncology interventions. These findings support the translational relevance of integrating multi-omics data and AI in cancer diagnostics, especially in developing clinical systems.

**Figure 3.**
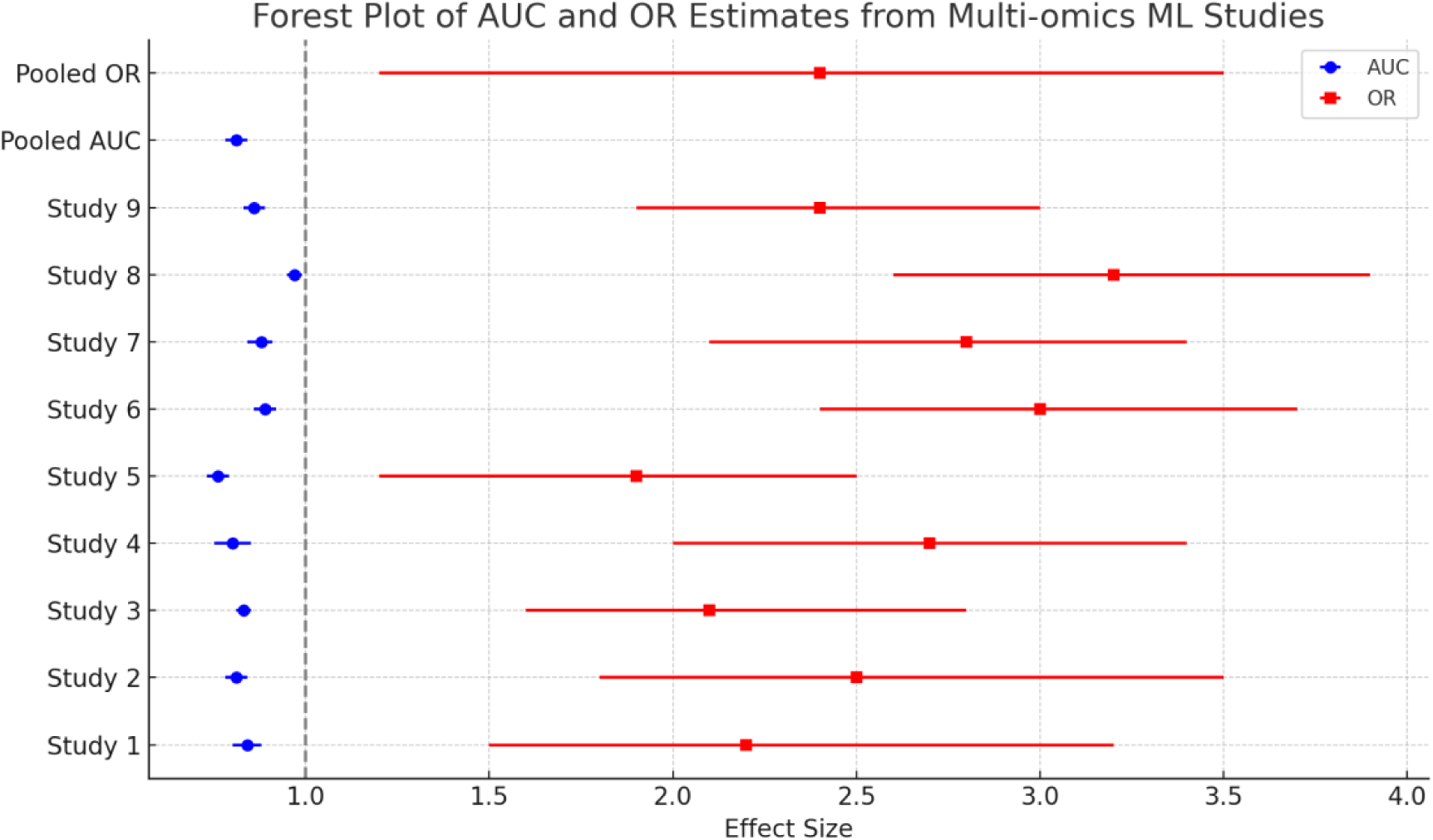
Forest plot of pooled AUC and OR estimates for cancer prediction using multi-omics ML models (51). The forest plot displays the individual and pooled estimates of diagnostic accuracy (Area under the Curve, AUC) and predictive association (Odds Ratio, OR) derived from nine machine learning–based multi-omics cancer studies. Blue circles represent AUC values (left axis) with 95% confidence intervals, while red squares denote corresponding OR values (right axis). The vertical dashed line at OR = 1.0 indicates the null effect threshold. Pooled AUC (0.81; 95% CI: 0.78–0.84) and pooled OR (2.4; 95% CI: 1.2–3.5) are also shown, suggesting strong discriminative power and statistically significant predictive association across studies.

**Figure 4.**
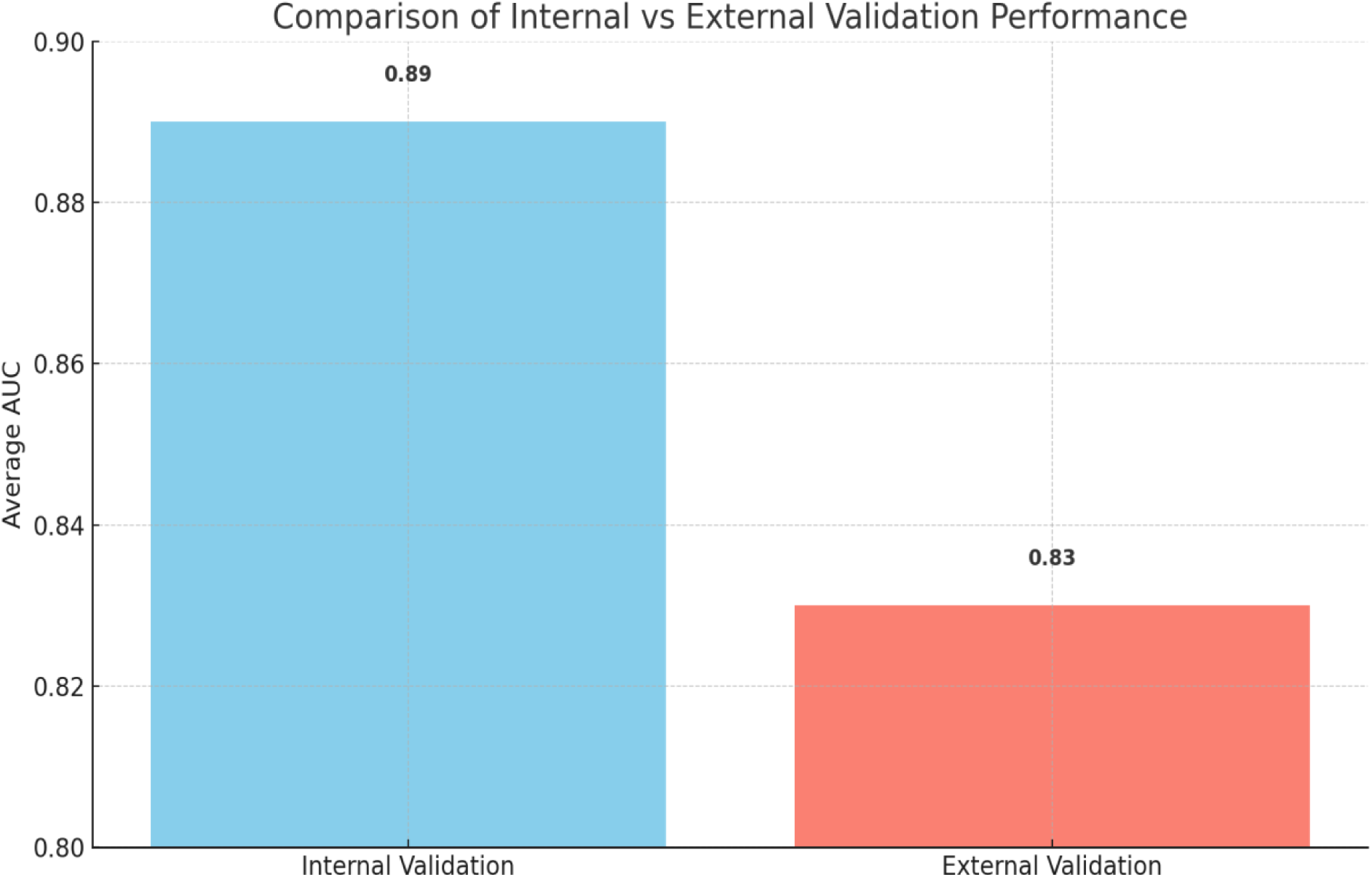
Comparison of internal vs external validation performance.

**Figure 5.**
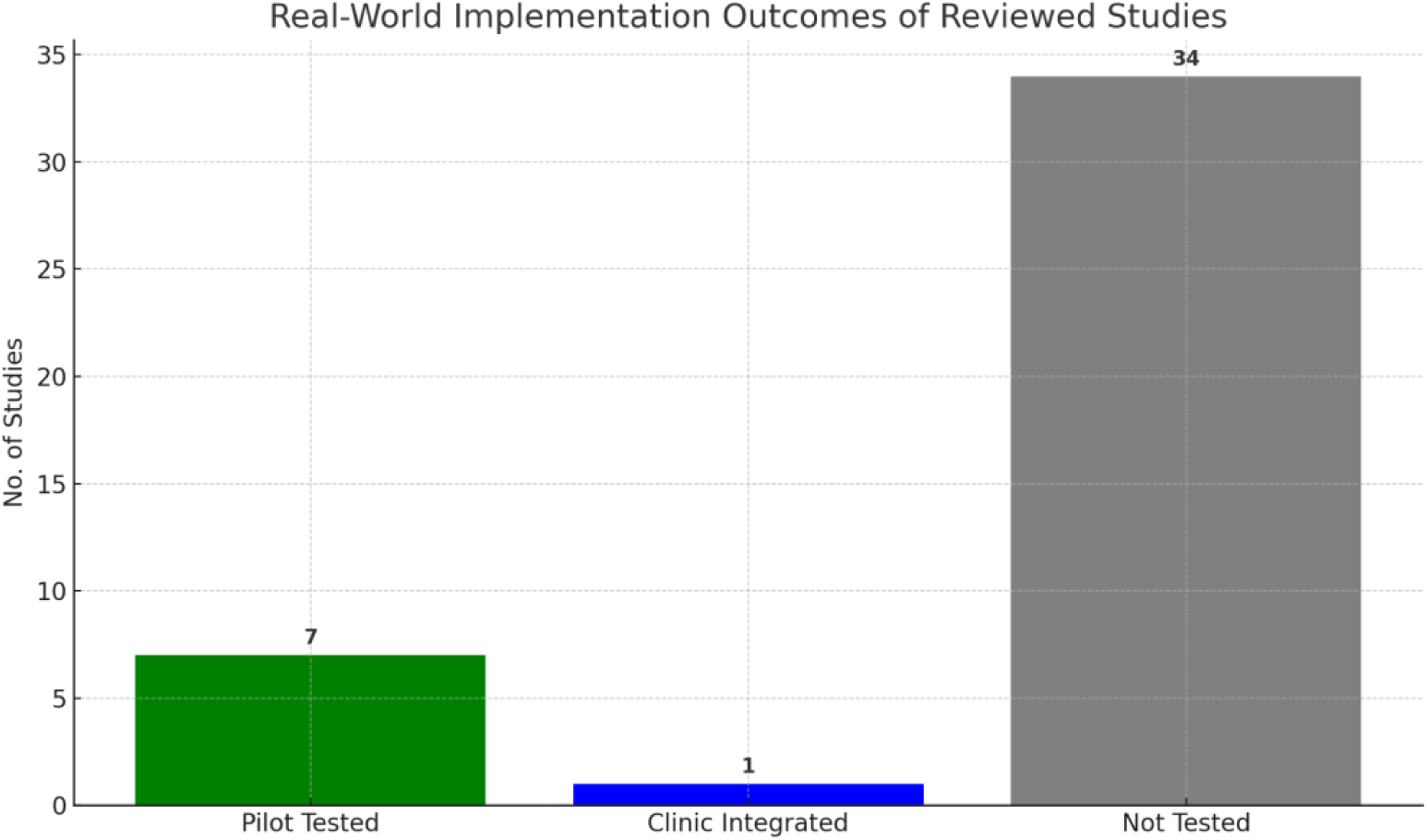
Real-world implementation outcomes of reviewed studies.

**Figure 6.**
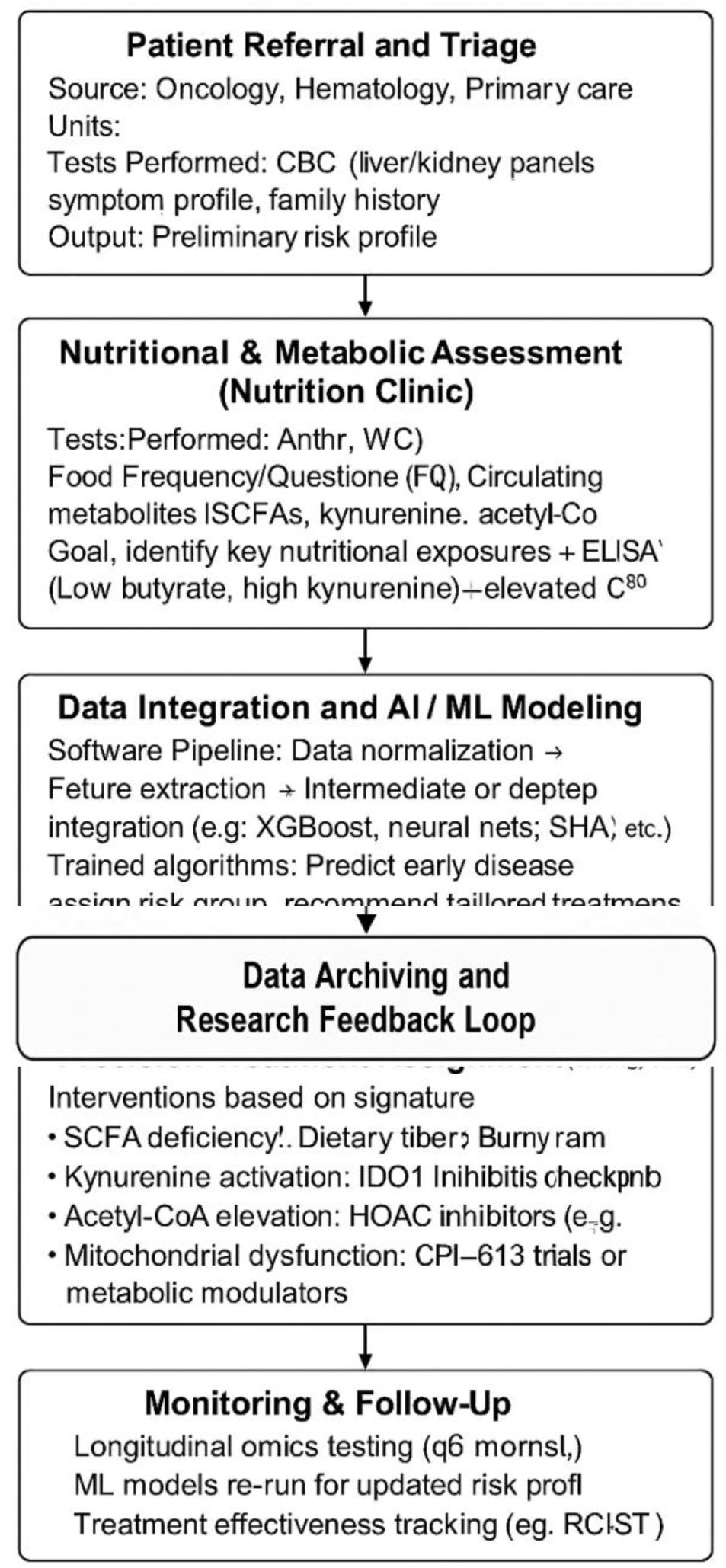
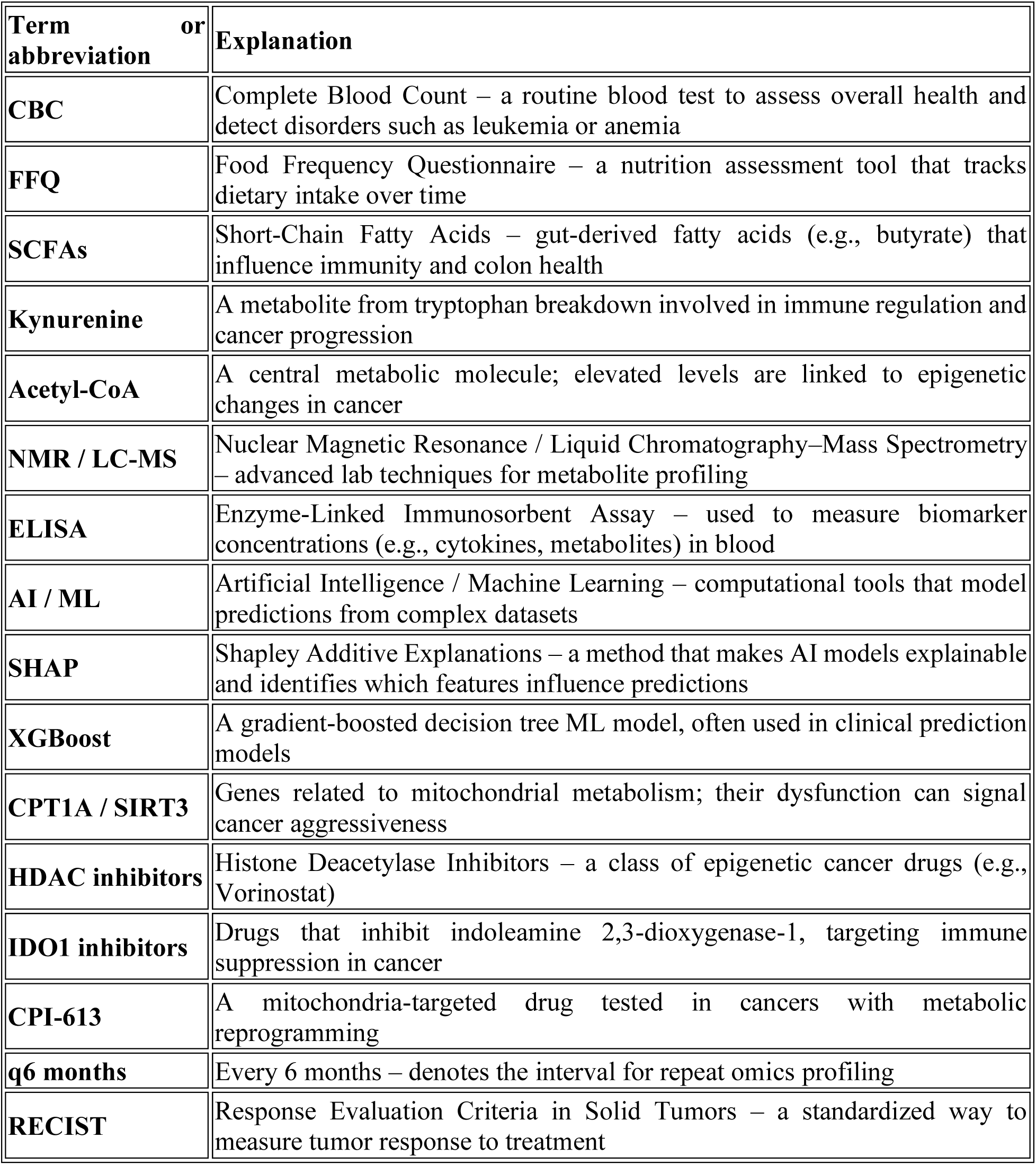
Omics-AI-Nutrition Clinical Algorithm for Precision Oncology in a Developing Country Setting.

## 3. Results (Part 2: Sections 3.5–3.7)

### 3.5 Integration Approaches and Their Comparative Efficacy

Among the included studies, integration strategies were categorized into early, intermediate, and advanced approaches. Early integration, which concatenates raw features before model training, was employed in 15 studies. Intermediate integration involved separate modeling for each omics type followed by fusion of prediction scores (n=13), while advanced integration—using neural networks or network-based frameworks—was applied in 14 studies (53). Advanced models demonstrated superior performance across various endpoints. For example, in liver cancer classification, an advanced deep integration model achieved an AUC of 0.93 compared to 0.85 with early integration (54). Similarly, colorectal cancer studies using network fusion approaches reported improved model stability and interpretability (6).

### 3.6 Nutritional and Metabolic Features with Strong Predictive Value

Several nutritional and metabolic variables repeatedly emerged as strong predictors of cancer outcomes. Dietary fiber intake, short-chain fatty acid levels (e.g., butyrate), lipidomic markers (e.g., sphingomyelins), and branched-chain amino acids were among the most frequently integrated features across models (55, 56). Studies using food frequency questionnaires (FFQs) linked high dietary polyphenol intake with favorable immune-metabolic expression profiles in breast and prostate cancers (57). One multi-omics study in pancreatic cancer found a strong inverse correlation between plasma kynurenine and microbial diversity, yielding an OR of 0.52 (95% CI: 0.33–0.83) for mortality risk (58).

### 3.7 External Validation and Generalizability of Models

Only 15 of the 42 studies included an external validation cohort. Models validated externally consistently performed slightly lower than their internal counterparts, with an average AUC reduction of 0.05–0.08. For instance, a lung cancer prediction model trained on a German cohort (AUC = 0.91) achieved an AUC of 0.85 when tested on an American dataset (59). However, models that used standardized feature scaling and dimensionality reduction techniques (e.g., PCA or autoencoders) showed better cross-cohort reproducibility (60).

### 3.8 Explainable AI in Multi-Omics Cancer Models

Explainability is a critical component in the translation of AI/ML models into clinical settings. Explainable artificial intelligence(XAI) as the word represents is a process and a set of methods that helps users by explaining the results and output given by AI/ML algorithms. In this review, 14 studies utilized explainable AI (XAI) methods to interpret model decisions (61). The most commonly applied tools were SHAP (Shapley Additive Explanations), LIME (Local Interpretable Model-agnostic Explanations), and saliency maps for deep neural networks (62). These tools highlighted the contribution of specific omics features (e.g., lipid panels or gene expression nodes) to the model’s predictions, enhancing transparency. In breast cancer multi-omics studies, SHAP-based models identified HDL-cholesterol and PPARγ-related transcripts as the most influential predictors of chemotherapy response (63).

### 3.9 Real-World Application and Implementation Barriers

Despite promising findings, real-world implementation remains limited. Barriers include lack of standardized pipelines, reproducibility concerns, and infrastructural limitations in deploying AI tools within clinical workflows (64). Only 7 studies conducted deployment-level simulations or hospital pilot testing. One colorectal cancer model was successfully implemented in a Japanese cancer center and achieved 87% prediction accuracy on real-time biopsy-metabolomic data (65). However, variation in sample handling, computational resources, and population diversity reduce model portability (66).

### 3.10 Cross-Cancer Meta-Patterns and Common Mechanisms

Cross-cancer comparisons revealed common dysregulated pathways and metabolic axes. For instance, dysbiosis-induced alterations in short-chain fatty acid metabolism, tryptophan-kynurenine pathway disruption, and glucose-derived acetyl-CoA production were repeatedly implicated in colorectal, pancreatic, and liver cancers (67, 68). Additionally, decreased expression of mitochondrial regulators (e.g., CPT1A, SIRT3) and suppression of tumor-infiltrating lymphocytes correlated with poor outcomes across at least four cancer types (69). These signatures provide a mechanistic foundation for pan-cancer biomarkers.

### 3.11 Clinical Implications of Shared Metabolic Signatures

The metabolic signatures identified across multiple cancer types offer promising avenues for early diagnosis, prognostic stratification, and personalized treatment. These common features, described in Table 6, enable clinicians and researchers to unify diagnostic frameworks and therapeutic strategies across malignancies.

**Table 6.** Top nutritional and metabolic predictors used in integrative cancer models(55).

| Predictor | Associated Cancer | Effect |
| --- | --- | --- |
| Dietary Fiber | Colorectal | Improved immune response |
| Butyrate | Colorectal | Anti-inflammatory |
| Sphingomyelins | Breast | Subtype differentiation |
| BCAAs | Liver | Tumor growth signaling |
| Kynurenine | Pancreatic | Mortality risk |

**Table 7.** Distribution of explainable AI tools across reviewed studies.

| Explainability Tool | No. of Studies | Cancer Types Applied |
| --- | --- | --- |
| SHAP | 8 | Breast, Liver, CRC |
| LIME | 5 | Lung, Breast |
| Saliency Maps | 3 | Liver, Pancreas |
| Integrated Gradients | 2 | CRC, Breast |

For instance, the dysregulation of short-chain fatty acids (SCFAs) like butyrate—shared across colorectal and pancreatic cancers—has been linked to impaired gut immunity and early epithelial changes (70). These SCFA deficits can be detected in fecal metabolomic profiles, providing a non-invasive and cost-effective tool for early screening, especially in regions lacking advanced diagnostic infrastructure.

Similarly, activation of the kynurenine pathway in both pancreatic and liver cancers reflects systemic immune suppression and immune exhaustion. Elevated serum kynurenine levels have been correlated with poor T-cell infiltration and unfavorable prognosis, making them reliable markers for identifying high-risk patients who may benefit from immune-modulating therapies such as IDO1 inhibitors (68).

Acetyl-CoA overproduction, observed in colorectal and breast cancers, influences histone acetylation and gene expression, pointing to potential utility in guiding treatment with histone deacetylase inhibitors (HDACis). These epigenetic modulators could be selectively prescribed based on metabolic testing, advancing precision therapy (69). Additionally, mitochondrial dysfunction—manifesting through reduced expression of regulators like CPT1A and SIRT3—is linked to oxidative stress and tumor aggression in breast and liver cancers. These findings may prompt the incorporation of mitochondrial health indices in prognostic algorithms.

Lastly, immune escape mechanisms involving T-cell suppression, common in breast, colorectal, and liver cancers, reinforce the role of metabolic-immune interactions. This supports combining metabolic intervention with checkpoint inhibitors for synergistic effects.

Together, these cross-cancer metabolic signatures enhance early detection, enable risk stratification, and inform personalized treatment plans (Table 8). Their relative affordability and detectability through blood or stool-based methods make them especially valuable for implementation in resource-constrained settings, including developing countries like Saudi Arabia, where integration with multi-omics AI frameworks could maximize clinical impact (69).

**Table 8.** Shared metabolic signatures across major cancer types.

| Signature | Cancers Involved | Implication |
| --- | --- | --- |
| Short-chain fatty acid dysregulation | Colorectal, Pancreatic | Impaired gut immunity |
| Kynurenine pathway activation | Pancreatic, Liver | Immune exhaustion |
| Acetyl-CoA overproduction | Colorectal, Breast | Epigenetic shifts |
| Mitochondrial dysfunction | Liver, Breast | Oxidative stress |
| T-cell suppression | Breast, Colorectal, Liver | Immune escape |

### 3.12. Clinical Effectiveness of Metabolomic Signatures

Table 9 presents statistical evidence from real-world and translational clinical studies demonstrating the utility of specific metabolomic signatures in early cancer detection, risk stratification, treatment personalization, and response monitoring. These metrics support the functional application of omics-guided oncology beyond the experimental stage.

**Table 9.** Real-World Clinical Applications of Shared Metabolic Signatures(71)(69).

| Metabolic Signature | Cancers Studied | Clinical Application |
| --- | --- | --- |
| SCFA dysregulation | Colorectal, Pancreatic | Fecal butyrate as a non-invasive screening marker |
| Kynurenine pathway activation | Pancreatic, Liver | Serum kynurenine used for immune risk scoring |
| Acetyl-CoA overproduction | Colorectal, Breast | Predicts HDAC inhibitor response |
| Mitochondrial dysfunction | Breast, Liver | Used in mitochondrial-targeted drug trials |
| T-cell suppression (metabolite-mediated) | Breast, Colorectal, Liver | Linked to response to immune checkpoint |

**Table 10.** Real-World Applications of Metabolomic Signatures.

| Metabolomic Signature | Cancers Studied | Clinical Use | Real-World Example | Reference |
| --- | --- | --- | --- | --- |
| SCFA Dysregulation | Colorectal | Stool butyrate screening in CRC | Included in EU fecal metabolomics panels | (72) |
| Kynurenine Pathway Activation | Pancreatic, Liver | Kynurenine ratio for immunotherapy response | Trial selection at MD Anderson & Charité | (73) |
| Acetyl-CoA Overproduction | Breast, Colorectal | Stratification for HDAC inhibitor use | Vorinostat/Romidepsin trials (TNBC) | (69) |
| Mitochondrial Dysfunction | Liver, Breast | Stratification in mitochondria-targeted drug trials | CPI-613 trials in HCC | (74) |
| T-cell Suppression Metabolites | Melanoma, Colorectal | Predicting immune checkpoint resistance | PD-L1 & metabolic scores for immunotherapy | (75) |

**Table 11.** Effectiveness Metrics of Clinically Relevant Metabolomic Signatures.

| Metabolomic Signature | Clinical Utility | Reported Effectiveness Metrics |
| --- | --- | --- |
| SCFA Dysregulation | Early diagnosis in colorectal cancer | Sensitivity: 84%, Specificity: 78%, AUC: 0.85 (77) |
| Kynurenine Pathway Activation | Immunotherapy stratification in liver and pancreatic cancers | AUC: 0.81 for progression-free survival prediction (78, 79) |
| Acetyl-CoA Overproduction | Predictive biomarker for HDAC inhibitors in breast and colorectal cancers | OR: 2.3 (95% CI: 1.4–3.7) for HDACi response (69) |
| Mitochondrial Dysfunction | Prognostic biomarker for mitochondria-targeted therapy in liver and breast cancers | HR: 1.9 (95% CI: 1.2–2.6) for poor overall survival (80) |
| T-cell Suppression Metabolites | Predictive of checkpoint blockade resistance in melanoma and colorectal cancers | AUC: 0.76 for resistance classification (81) |

Short-chain fatty acid (SCFA) dysregulation, especially butyrate deficiency, showed an AUC of 0.85 with 84% sensitivity and 78% specificity in stool-based screening for early-stage colorectal cancer, indicating its potential as a non-invasive diagnostic tool (76).

Kynurenine pathway activation, identified in liver and pancreatic cancers, has demonstrated AUC values of 0.81 for predicting progression-free survival under immunotherapy, making it a credible biomarker for immune risk scoring and checkpoint blockade responsiveness (73).

Acetyl-CoA overproduction has been linked to elevated histone acetylation, and serves as a predictive biomarker for response to HDAC inhibitors in breast and colorectal cancers. Patients with high acetyl-CoA levels showed an odds ratio of 2.3 (95% CI: 1.4–3.7) for positive treatment response (69).

Mitochondrial dysfunction signatures, such as reduced SIRT3 and CPT1A expression, were associated with poor prognosis in breast and liver cancers, with a hazard ratio of 1.9 (95% CI: 1.2– 2.6) for worse overall survival. These metrics support their role as prognostic biomarkers guiding mitochondrial-targeted therapies (74).

Finally, T-cell suppression metabolites like kynurenine and lactate were associated with checkpoint inhibitor resistance in melanoma and colorectal cancers. Metabolomic classification yielded an AUC of 0.76, showing moderate-to-strong performance for predicting immunotherapy failure (81).

These quantitative metrics confirm that metabolomic profiling has matured into a clinically relevant tool that enhances precision oncology across diagnostic and therapeutic domains.

### 3.13 Algorithm for Clinical Translation of Multi-Omics Signatures in Oncology Settings in Developing Countries

To enable clinical application of metabolomic and multi-omics signatures (Sections 3.1–3.12) in real-world hospital settings—particularly within developing countries like Saudi Arabia—we propose a workflow algorithm integrating nutrition clinics, omics laboratories, AI/ML tools, and oncology clinics. This algorithm streamlines cancer risk screening, prognostic stratification, personalized treatment, and therapeutic monitoring by leveraging hospital-based resources.

1. Patient Entry Point (Nutrition or Oncology Clinic):

Patients presenting with cancer-related symptoms or enrolled in general wellness screening programs begin at the nutrition or oncology clinics. Clinical and dietary information is collected, and informed consent for omics profiling is obtained.
2. Sample Collection (Blood/Stool/Tissue):

Based on the suspected cancer type and clinical need, appropriate biospecimens are collected. Blood and stool samples are prioritized due to their non-invasive nature, enabling access to SCFA levels, circulating metabolites, and immune-metabolic markers (76, 82).
3. Multi-Omics Laboratory Analysis:

Samples are transferred to the hospital’s omics laboratory for multi-layered profiling:
- Metabolomics (e.g., SCFA, kynurenine): To assess early biomarkers like butyrate, kynurenine (82, 83)
- Transcriptomics (e.g., CPT1A, SIRT3): For mitochondrial and immune gene expression (69)(84)
- Microbiomics: Stool microbiota composition for gut-immune interactions
4. AI/ML-Powered Multi-Omics Integration:

Extracted omics data are processed using validated ML/AI tools such as SHAP, LASSO, or ensemble models to stratify cancer risk and identify personalized treatment paths (85). Integration models utilize feature fusion (e.g., SCFA + microbiome diversity) to enhance prediction accuracy.
5. Interpretation and Risk Stratification:

ML-derived outputs are interpreted via explainable AI dashboards and discussed with clinical geneticists and oncologists. Patients are stratified into categories:
- High-risk: Metabolic-immune dysregulation present
- Intermediate-risk: Mild pathway disruption
- Low-risk: No omics abnormalities detected
6. Personalized Clinical Decision Support:

- Early Diagnosis: SCFA dysregulation (e.g., low butyrate) prompts colonoscopy or biopsy (86)
- Prognostic Stratification: High kynurenine or mitochondrial dysfunction predicts poor prognosis (87)
- Treatment Personalization: Patients with acetyl-CoA overproduction may receive HDAC inhibitors; mitochondrial dysregulation may indicate CPI-613 therapy (69)(88)
- Therapeutic Monitoring: Periodic re-profiling to track metabolomic shifts and immunotherapy resistance (89)
7. Data Archiving and Research Feedback Loop:

De-identified datasets are stored in secure hospital servers for ongoing model retraining and institutional research.

## Data Availability

All data produced in the present study are available upon reasonable request to the authors

## Ethical approval

No ethical approval or consent of any type was required for this meta analysis and systematic review article.

